# Evaluating Metformin Efficacy in ALS Using Real-World Data: A Causal Inference Approach

**DOI:** 10.64898/2026.03.11.26347714

**Authors:** Julia A. Geller, Alex Berger, Matteo Locatelli, Senda Ajroud-Driss, Tawfiq Al-Lahham, Ximena Arcila-Londono, Federica Cerri, Vivian Drory, Mehdi Ghasemi, Marc Gotkine, Kelly G. Gwathmey, Ghazala Hayat, Terry Heiman-Patterson, Christian Lunetta, Nicholas Olney, Jefferey Rosenfeld, David Walk, James Wymer, Alexander V. Sherman, Marie-Abele Bind

## Abstract

**Background and Objectives:** Observational data can provide evidence of treatment effects in a low-burden, cost-effective manner. Using a causal inference framework to address selection bias, we illustrate the approach in ALS by examining previously researched associations, baseline function and sex, and evaluating metformin’s survival benefits.

**Methods:** Data are from the ALS Natural History Study collected in the United States, Israel, and Italy from 2015 to 2025. We employ propensity score matching, two approaches to address loss to follow-up (i.e. a naive exclusion and principal stratification), and randomization-based inference to evaluate difference in 18-month restricted mean survival time.

**Results:** For the function investigation, estimated survival is 16.39, 16.14, 16.09, and 16.24 months for the high ALSFRS-R group and 14.04, 14.42, 14.72, and 14.60 months for the low ALSFRS-R group using the non imputed and imputed datasets 1-3, respectively (p<0.0001, N=450; p<0.00001, N=1,485; p<0.00001, N=1,492; p<0.00001, N=1,600). Using principal stratification, estimated survival is 16.38, 16.14, 16.09, and 16.23 months for the high ALSFRS-R group and 14.04, 14.42, 14.72, and 14.59 months for the low ALSFRS-R group with the non imputed and imputed datasets 1-3, respectively (p<0.0001, N=449; p<0.00001, N=1,483; p< 0.00001, N=1,491; p<0.00001, N=1,596).

For the sex investigation, estimated survival is 15.20, 15.16, 15.20, and 15.17 months for females and 15.41, 14.84, 14.91, and 14.97 months for males using the non imputed and imputed datasets 1-3, respectively (p≈0.594, N=612; p≈0.155, N=2,081; p≈0.200, N=2,104; p≈0.359, N=2,111). Using principal stratification, estimated survival is 15.20, 15.16, 15.20, and 15.17 months for females and 15.41, 14.83, 14.91, and 14.97 months for males with the non imputed and imputed datasets 1-3, respectively (p≈0.598, N=612; p≈0.146, N=2,079; p≈0.195, N=2,102; p≈0.364, N=2,110).

For the metformin investigation, estimated survival is 14.77, 14.87, and 14.57 months for metformin users and 14.59, 14.43, and 13.97 months for non metformin users using imputed datasets 1-3, respectively (p≈0.391, N=289; p≈0.257, N=279; p≈0.191, N=301). Using principal stratification, estimated survival is 14.75, 14.85, and 14.55 months for metformin users and 14.68, 14.58, and 13.97 months for non metformin users with imputed datasets 1-3, respectively (p≈0.458, N=286; p≈0.351, N=275; p≈0.201, N=300).

**Discussion:** We reject the sharp null hypothesis for the function investigation and fail to reject it for the sex and metformin investigations.

**Trial Registration Information:** ClinicalTrials.gov Identifier: NCT05966038.

## 1 INTRODUCTION

Amyotrophic Lateral Sclerosis (ALS) is a fatal neurodegenerative disease with few successful phase III trials, where trial design balances complex ethical and practical considerations. Although randomized controlled trials (RCTs) are considered the gold-standard for estimating causal effects, they face substantial challenges in ALS. Heterogeneity in disease progression between people living with ALS may obscure treatment effects (Kiernan *et al*., 2021). The optimal clinical trial duration for ALS remains unclear. Most phase II trials employ a 6-month placebo-controlled period.

However, a combined analysis of double-blind and open label extension data for tofersen in *Superoxide Dismutase 1* (*SOD1*) ALS showed a smaller decline in ALSFRS-R at 12 months (Miller *et al*., 2022), raising questions about whether traditional durations are sufficient for outcomes with limited sensitivity and treatments that may have delayed effects. However, extending trial durations may not be feasible given substantial costs. RCTs impose burden on patients by requiring them to attend study-specific visits and present ethical concerns about withholding treatments from placebo groups (Kiernan *et al*., 2021), (Colli *et al*., 2014). Platform trials with a master protocol and shared control-arm to evaluate multiple agents may help alleviate ethical concerns, administrative delays, and some financial barriers (Kiernan *et al*., 2021), as seen in the Healey ALS Platform Trial (Quintana *et al*., 2023). Additionally, observational approaches may advance the pace of therapeutic evaluation in ALS by providing real-world evidence in a cost-effective, low-burden manner to evaluate new hypotheses and confirm existing findings. By recording a wide range of clinically meaningful data, observational studies enable the study of multiple hypotheses over the entire collection period. Furthermore, drug repurposing may be advantageous for ALS where there is an urgent need for effective treatments by using an established safety profile to advance phase II assessments and speed up trial timelines (Kiernan *et al*., 2021).

Observational analysis of metformin, an antidiabetic drug increasingly studied in neurological conditions, can provide evidence of its effects in ALS. Metformin is a first-line treatment for type 2 diabetes (Kruczkowska *et al*., 2025; Loan *et al*., 2024) with phase II trials for mild cognitive impairment (ClinicalTrials.gov, 2025b), Parkinson’s disease (ClinicalTrials.gov, 2025a), Huntington’s disease (ClinicalTrials.gov, 2025d), and ALS-frontotemporal dementia (ALS-FTD) (ClinicalTrials.gov, 2025c). The drug has emerged as a promising therapy for neurodegenerative diseases as it operates through pathways implicated in cellular energy metabolism and autophagy, neuroinflammation, protein aggregation, and mitochondrial dysfunction, common pathological features of neurodegenerative conditions. Preclinical studies suggest that metformin may have varied effects based on genetic variants of ALS and sex. A study in *C9orf72* ALS-FTD mice suggested that metformin improves anxiety-like behavior and reduces neuroinflammation and motor neuron loss (Zu *et al*., 2020), while another study in the *SOD1* ALS mouse model failed to reject the null hypothesis for effects on disease onset or survival in male mice but reported that metformin was associated with faster symptom onset in female mice in a dose-dependent manner (Kaneb *et al*., 2011). Associations between antidiabetic medications and ALS risk have been reported (Diekmann *et al*., 2020; Mariosa *et al*., 2020), but the role of metformin was not specifically studied. Furthermore, an analysis of reported adverse events with the United States Food and Drug Administration (FDA) MedWatch data found that metformin was associated with a reduced risk of ALS (Lehrer and Rheinstein, 2024). Overall, the survival benefit of metformin for people living with ALS across all disease subtypes remains understudied.

Pre-clinical studies of metformin have been limited to the *C9orf72* and *SOD1* variants and are not directly applicable to other genetic variants or sporadic ALS. Additionally, studies examining antidiabetic use and ALS risk estimated associations rather than causal effects. Although there is an active phase II trial of metformin in people living with ALS, the study is limited to the *C9orf72* genetic variant (ClinicalTrials.gov, 2025c). To address these limitations, we employ the Rubin Causal Model (Rosenbaum and Rubin, 1983; Imbens and Rubin, 2015; Rubin, 2008) within a four-stage causal pipeline (Bind and Rubin, 2017) to assess causal questions. First, we evaluate two previously-researched hypotheses in ALS to illustrate the approach, the association of baseline function and sex with survival. Then, we estimate the causal effects of a novel treatment, metformin, on survival in a broad cohort of people living with ALS. We are interested in the following questions:

1. *Is baseline function positively associated with 18-month survival?*
2. *Is sex associated with 18-month survival?*
3. *Does metformin usage have a positive effect on 18-month survival?*

## 2 METHODS

### 2.1 The ALS Natural History Study

This study includes 2,727 people living with ALS from the *ALS Natural History Study* (ALS NHS). The ALS NHS is an observational study with seventeen multidisciplinary clinical sites across the United States, Italy, and Israel. Data are accessed through NeuroBank™, a clinical research platform developed by Mass General Brigham (ALS/MND Natural History Consortium, 2026). We consider the time period from the beginning of recruitment to date of data access, from June 15^th^, 2015 to October 20^th^, 2025. During standard care visits, typically every three months, trained health professionals record participant demographics, collect electronic health record information, and conduct validated assessments. Individuals must receive care at a member site and have an ALS diagnosis to participate. In our analysis, we exclude participants with Primary Lateral Sclerosis (PLS) due to notably longer survival from other ALS phenotypes (Masrori and Van Damme, 2020).

Demographic information includes sex, age at symptom onset, age at diagnosis, race, ethnicity, and BMI. Disease-specific characteristics include El Escorial diagnosis, phenotype, site of symptom onset, diagnostic delay (time from symptom onset to diagnosis), enrollment delay (time from diagnosis to baseline visit), disease duration (time from symptom onset to baseline visit), maximum vital capacity (the highest percent predicted value among slow vital capacity and forced vital capacity measured in upright and supine positions), and genetic variants related of ALS. Function is measured by the ALS Functional Rating Scale-Revised total score (ALSFRS-R), a validated questionnaire that scores physical function across bulbar, fine motor, gross motor, and respiratory domains where 48 is the highest and 0 the lowest level of function (Cedarbaum *et al*., 1999). To account for FDA-approved and the most common medications taken by people living with ALS in the ALS NHS, we include reported history of riluzole, radicava, gabapentin, baclofen, nuedexta, and omeprazole. We also include the reported history of the most common concurrent conditions among people living with ALS in the ALS NHS: hypertension, hyperlipidemia, depression, sleep apnea, asthma, anxiety, obesity, hypothyroidism, and allergic rhinitis. For the metformin investigation, we consider reported history of metformin use and reported history of pre-diabetes and diabetes, the diagnoses metformin is primarily prescribed for. Survival, measured in months from baseline visit to last visit, loss to follow up date, or death, was selected as the outcome due to its clinical relevance and prior use in demonstrating the efficacy of riluzole in ALS (Bensimon *et al*., 1994).

### 2.2 Causal Inference Framework

Causal inference methods provide a framework to evaluate treatment effects using real-world data. The Rubin Causal Model (RCM) was successfully employed in a variety of environmental, epidemiology, and criminology literature to assess causal effects (Sommer *et al*., 2022, 2018, 2020) and can be adapted to study ALS-related hypotheses. The key challenge of causal inference is one cannot observe how the same individual would respond to a different exposure under equal conditions (Imbens and Rubin, 2015). Randomization ensures that a difference between groups is present by chance, so repeated studies produce estimates that on average approximate the treatment effect. However, observational data are inherently non-randomized and could reflect selection bias and influence from confounders (Rubin, 1974). The RCM aims to minimize external influences by constructing similar treatment and control groups, facilitating “apples to apples” comparisons and enabling the estimation of unbiased estimates of causal effects (Sommer *et al*., 2018) under transparent assumptions. In the following sections, we employ the RCM in a causal ALS setting (Bind and Rubin, 2017).

### 2.3 Stage 1: Conceptual Stage

The process begins with the conceptual stage, where the goal is to precisely define the question of interest and formulate a hypothetical randomized experiment. During this stage, the study population, treatment or intervention, outcomes, and time period are defined (Bind and Rubin, 2017). We define the treatment assignment *W*_*i*_ by:

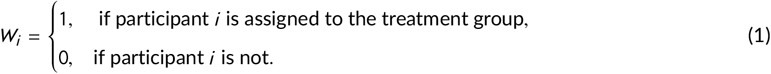

The RCM describes the potential outcomes, where each participant can have two possible outcomes. For participant *i*, when assigned to the treatment group (*W*_*i*_ = 1) the outcome is *Y*_*i*_ (*W*_*i*_ = 1), and when assigned to the control group (*W*_*i*_ = 0) the outcome is *Y*_*i*_ (*W*_*i*_ = 0). Only one outcome is observed, defined as:

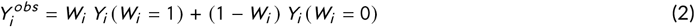

The Stable Unit-Treatment Value Assumption (SUTVA) must hold to employ the RCM (Rubin, 2008), which in our study asserts that each patient receives one version of the treatment and that there is no interference between patients.

### 2.4 Stage 2: Design Stage

The aim of the design stage is to construct two groups with comparable distributions of confounding variables (Rubin, 1974, 2008; Rosenbaum and Rubin, 1983). We conduct this stage by matching participants on an estimated propensity score, which is the estimated probability of a participant *i* to be assigned to the treatment group given background covariates *X*_*i*_, i.e., *P* (*W*_*i*_ = 1|*X*_*i*_) (Rubin, 2008). By reducing differences in estimated propensity scores, individual differences in covariate distributions are equally reduced to create similar groups (Stuart, 2010). Participants with propensity scores outside overlapping score distributions between groups and all remaining unpaired participants were excluded (Stuart, 2010). We used logistic regression to estimate propensity scores and one-to-one matching. To avoid inadequate matches, propensity score differences within pairs had to be under a chosen threshold, known as the caliper.

We present group balance with Love plots and balancing quality is quantified with absolute standardized mean differences (SMDs). All SMDs must be between ≤ 0.25 for reliable estimates (Stuart, 2010), though in practice ≤ 0.1 is more commonly used. Background covariates for estimating propensity scores were selected by excluding covariates balanced pre-matching and iteratively including those imbalanced post-matching until the best balance was achieved. It should be noted that covariates used for propensity score estimation must be available at the chosen study start date and outcomes should not be viewed during this stage to minimize bias (e.g., (Bind and Rubin, 2017)) However, excluding variables associated with the outcome can subtly increase bias in causal estimates which may not be detected (Stuart, 2010). Details on covariates and calipers used for each investigation are described in the Appendix.

### 2.5 Stage 3: Analysis Stage

Next, the analysis can be performed to estimate causal effects. It is best practice to specify an analysis protocol *a priori* without viewing outcomes to avoid introducing bias (Rubin, 2008).

#### 2.5.1 Sharp Null Hypothesis

We evaluate the sharp null hypothesis stating that survival does not differ by assignment status for any participant *i*, expressed as:

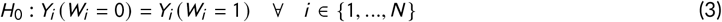

#### 2.5.2 Randomization-Based Inference

As the test statistic, we use the difference in restricted mean survival time between groups, where the restricted mean is defined by:

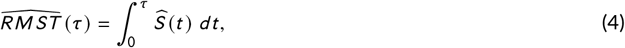

where 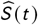 (*t*) is the kaplan-meier survival curve up to time point τ.

When conducting randomization-based inference, treatment assignments are permuted for *N*_*i ter*_ iterations, recomputing the test statistic at each step to construct the null randomization distribution. For a right-tailed test, the p-value is defined as the proportion of test statistics as or more extreme than the observed test statistic, formally expressed as 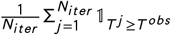 where 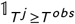 is 1 when *T* ^*j*^ ≥ *T* ^*obs*^ otherwise 0. For a two-sided test, the p-value *N*_*i t er*_ is defined by the absolute value of test statistics, where 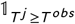 is 1 when |*T* ^*j*^ | ≥ |*T* ^*obs*^ | otherwise 0. In our analysis, *N*_*i ter*_ = 100, 000 and *τ* = 18 months.

#### 2.5.3 Study Dropout and Principal Stratification

Study dropout may lead to deceptive conclusions of causal effects if not adequately addressed (Rubin, 2006). For instance, consider a clinical trial where the treatment is effective, but induces side effects that increase the likelihood of dropout. Estimating effects among participants who completed the study may underestimate treatment effects and potentially lead to a type II error. Frangakis and Rubin argue that causal effects can be estimated for those who would never be lost to follow up under either group assignment (Frangakis and Rubin, 2002).

Principal stratification identifies a principal strata of participants who would never be lost to follow-up. Predictive models estimate participation status under the unobserved treatment assignment and those lost to follow-up under the observed or hypothetical assignment are excluded from analysis. In our study, we use logistic regression to model the probability of dropout in each group, defined as *P* (*S*_*i*_ (*W*_*i*_ = 1) = 1| *Z*_*i*_) to predict dropout if assigned to the treatment group and *P* (*S*_*i*_ (*W*_*i*_ = 0) = 1| *Z*_*i*_) to predict dropout in the control group, given covariates *Z*_*i*_ and the dropout indicator variable *S*_*i*_ for each participant *i*. Despite limited investigation of study dropout in ALS, a meta-analysis of clinical studies found that a better baseline ALSFRS-R was an independent, positive predictor of completing a study (Atassi *et al*., 2013). Based on this finding, we adopt the view that dropout is motivated by functional decline which may make it challenging to attend clinic visits. Consequently, we use the following variables (*Z*_*i*_) known to affect disease progression in ALS: body mass index (BMI), age at symptom onset, diagnostic delay, enrollment delay, baseline ALSFRS-R, bulbar symptom onset, and lower motor neuron predominant phenotype (Westeneng *et al*., 2018b).

#### 2.5.4 Sensitivity Analysis

There are 834 people living with ALS in the ALS NHS with data recorded for all variables of interest. To estimate causal effects in a larger sample, we repeated the aforementioned procedure with multiply imputed data. For each investigation, we use the *mice* package in R (Buuren and Groothuis-Oudshoorn, 2011) to generate three imputed datasets each imputed for five iterations. Multiclass categorical data were imputed with polytomous logistic regression, binary categorical data with logistic regression, and numerical data with predictive mean matching. Restricted imputation was applied to date variables to ensure realistic disease timelines in imputed data. Specifically, the symptom onset date was restricted between the date of birth and diagnosis date, diagnosis date between symptom onset date and baseline date, and the last visit date between the baseline date and October 20th, 2025. The loss to follow-up and mortality date fields were both imputed to be after the participant’s last visit date.

In addition to background covariates (*X*_*i*_), we include the following variables for imputation: age at baseline visit, date last known alive (defined as the latest of last visit, death, or loss to follow up dates), time from diagnosis to survival in months, ALSFRS-R and BMI collected during a follow-up visit that is at least a week after the baseline visit. These variables support more accurate imputation of age at symptom onset, age at diagnosis, loss to follow up date, mortality date, baseline ALSFRS-R, baseline BMI, and survival time. Derived variables were then recalculated from imputed values.

### 2.6 Stage 4: Summary Stage

Finally, results are interpreted and developed into recommendations. A small p-value suggests that the observed test statistic is rare under the sharp null hypothesis and a higher p-value does not necessarily indicate no effect. P-values are viewed as evidence towards a hypothesis rather than definitive statements of fact and strict reliance on a 0.05 threshold should be approached with caution (Wasserstein and and, 2016; Wasserstein *et al*., 2019).

### 2.7 Standard Protocol Approvals, Registrations, and Patient Consents

The written informed consent was obtained from all participants in the study.

### 2.8 Data Availability

The study protocol and the dataset used in the paper will be made available upon request to the corresponding author.

## 3 RESULTS

### 3.1 Characteristics of Study Participants

To assess the hypotheses, we use matched ALS NHS data constructed in the design stage using participants without missing data (N=834) and multiply imputed data (N=2,727). We describe participant characteristics for all investigations in the Appendix. Notably, the non-metformin group in matched imputed dataset 2 has more females than males), which is unusual as incidence and prevalence of ALS is greater in males (McCombe and Henderson, 2010). Matched participants had a median disease duration ranging from 13 to 16 months, 13 to 15 months, and 14 to 18 months for the function, sex, and metformin investigations respectively, while median survival from symptom onset in ALS has been observed to range from 20 to 48 months (Chiò *et al*., 2009).

### 3.2 Function Investigation

For the function investigation, treatment assignment *W*_*i*_ for participant *i* is defined by Equation (5), where the assignment threshold (37) is the median ALSFRS-R for all people living with ALS in the ALS NHS who had an ALSFRS-R collected at baseline.

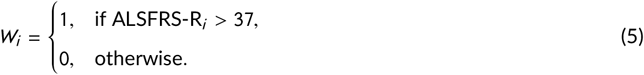

To evaluate the sharp null hypothesis, we conduct a test where the alternative hypothesis indicates a positive association between baseline ALSFRS-R and 18-month survival. Post-matching covariate SMDs are < 0.1 for nearly all covariates and the data is considered well balanced which is shown in the Appendix.

Using non-imputed data, estimated survival time was 16.39 (high ALSFRS-R group) and 14.04 months (low ALSFRS-R group) for individuals never lost to follow-up (p<0.0001; N=450) and 16.38 (high ALSFRS-R group) and 14.04 (low ALSFRS-R group) months using principal stratification (p<0.0001; N=449). Imputed dataset 1 showed 16.14 (high ALSFRS-R group) and 14.42 months (low ALSFRS-R group) for those never lost to follow-up (p<0.00001; N=1,485) and 16.14 (high ALSFRS-R group) and 14.42 months (low ALSFRS-R group) using principal stratification (p<0.00001; N=1,483). Imputed dataset 2 showed 16.09 (high ALSFRS-R group) and 14.72 months (low ALSFRS-R group) for those never lost to follow-up (p<0.00001; N=1,492) and 16.09 (high ALSFRS-R group) and 14.72 months (low ALSFRS-R group) using principal stratification (p<0.00001; N=1,491). Imputed dataset 3 showed 16.24 (high ALSFRS-R group) and 14.60 months for those never lost to follow-up (p<0.00001; N=1,600) and 16.23 (high ALSFRS-R group) and 14.59 months (low ALSFRS-R group) using principal stratification (p<0.00001; N=1,596).

Notably, difference estimates using imputed matched data across both dropout strategies were equivalent, with few individuals identified as lost to follow up under the hypothetical assignment using principal stratification. We therefore would like to study influences of study dropout more closely to ensure accuracy of principal strata models and better estimate treatment effects in the presence of censoring. Null randomization distributions for all analyses in the function investigation appeared to follow gaussian distributions centered at zero and can be found in the Appendix.

**TABLE 1.** Results of the function investigation. Estimated 18-month restricted mean survival time in months using 100,000 randomization iterations.

| Dataset (N Matched) | Dropout Strategy | High Function RMST (months) | Low Function RMST (months) | ΔRMST | P-value |
| --- | --- | --- | --- | --- | --- |
| Non-imputed (N=480) | Exclude (N=450) | 16.39 | 14.04 | 2.35 | $< 1e-04$ |
| | Never lost to follow-up (N=449) | 16.38 | 14.04 | 2.34 | $< 1e-04$ |
| First imputed (N=1,606) | Exclude (N=1,485) | 16.14 | 14.42 | 1.72 | $< 1e-04$ |
| | Never lost to follow-up (N=1,483) | 16.14 | 14.42 | 1.72 | $< 1e-04$ |
| Second imputed (N=1,610) | Exclude (N=1,492) | 16.09 | 14.72 | 1.37 | $< 1e-04$ |
| | Never lost to follow-up (N=1,491) | 16.09 | 14.72 | 1.37 | $< 1e-04$ |
| Third imputed (N=1,736) | Exclude (N=1,600) | 16.24 | 14.60 | 1.64 | $< 1e-04$ |
| | Never lost to follow-up (N=1,596) | 16.23 | 14.59 | 1.64 | $< 1e-04$ |

### 3.3 Sex Investigation

In the sex investigation, group assignment is determined by Equation (6)

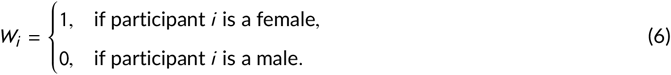

We employ a two-sided test to evaluate the sharp null hypothesis where the alternative hypothesis indicates an association between sex and 18-month survival. Love plots can be found in the Appendix. Across all datasets, the majority of covariate SMDs are < 0.1 post-matching with only two variables slightly above so the matching is successful.

Using non-imputed data, estimated survival time was 15.20 (females) and 15.41 months (males) for individuals never lost to follow-up (p≈0.594; N=612) and 15.20 (females) and 15.41 months (males) using principal stratification (p≈0.598; N=612). Imputed dataset 1 showed 15.16 (females) and 14.84 months (males) for those never lost to follow-up (p≈0.155; N=2,081) and 15.16 (females) and 14.83 months (males) using principal stratification (p≈0.146; N=2,079). Imputed dataset 2 showed 15.20 (females) and 14.91 months (males) for those never lost to follow-up (p≈0.200; N=2,104) and 15.20 (females) and 14.91 months (males) using principal stratification (p≈0.195; N=2,102). Imputed dataset 3 showed 15.17 (females) and 14.97 (males) months for those never lost to follow-up (p≈0.359; N=2,111) and 15.17 (females) and 14.97 (males) months using principal stratification (p≈0.364; N=2,110). As seen in the function investigation, few individuals were identified as lost to follow up under the hypothetical assignment and further investigation of dropout mechanisms in ALS may be necessary. The null randomization distributions for the sex investigations are shown in the Appendix and appeared to follow gaussian distributions.

### 3.4 Metformin Investigation

For the metformin investigation, treatment assignment *W*_*i*_ for participant *i* is defined by Equation (7).

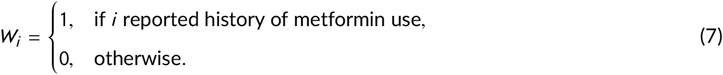

**TABLE 2.** Results of the sex investigation. Estimated 18-month restricted mean survival time in months using 100,000 randomization iterations.

| Dataset (N Matched) | Dropout Strategy | Female RMST (months) | Male RMST (months) | ΔRMST | P-value |
| --- | --- | --- | --- | --- | --- |
| Non-imputed (N=650) | Exclude (N=612) | 15.20 | 15.41 | -0.21 | 0.594 |
|  | Never lost to follow-up (N=612) | 15.20 | 15.41 | -0.21 | 0.598 |
| First imputed (N=2,256) | Exclude (N=2,081) | 15.16 | 14.84 | 0.32 | 0.155 |
|  | Never lost to follow-up (N=2,079) | 15.16 | 14.83 | 0.33 | 0.146 |
| Second imputed (N= 2,274) | Exclude (N=2,104) | 15.20 | 14.91 | 0.29 | 0.200 |
|  | Never lost to follow-up (N=2,102) | 15.20 | 14.91 | 0.29 | 0.195 |
| Third imputed (N=2,280) | Exclude (N=2,111) | 15.17 | 14.97 | 0.20 | 0.359 |
|  | Never lost to follow-up (N=2,110) | 15.17 | 14.97 | 0.20 | 0.364 |

To evaluate the sharp null hypothesis, we conduct randomization-based inference with a right-tailed test where the alternative hypothesis indicates a positive effect of metformin on 18-month survival. Post-matching covariate balance is poor in non-imputed data, with 5 covariates exhibiting SMDs > 0.2 post-matching. Balance in imputed data is substantially improved, with the majority of covariates achieving SMDs < 0.1 post-matching and only a few covariates slightly above 0.1 (Figures 1b–1d). We therefore proceeded with the metformin analysis only using imputed matched data.

**FIGURE 1.**
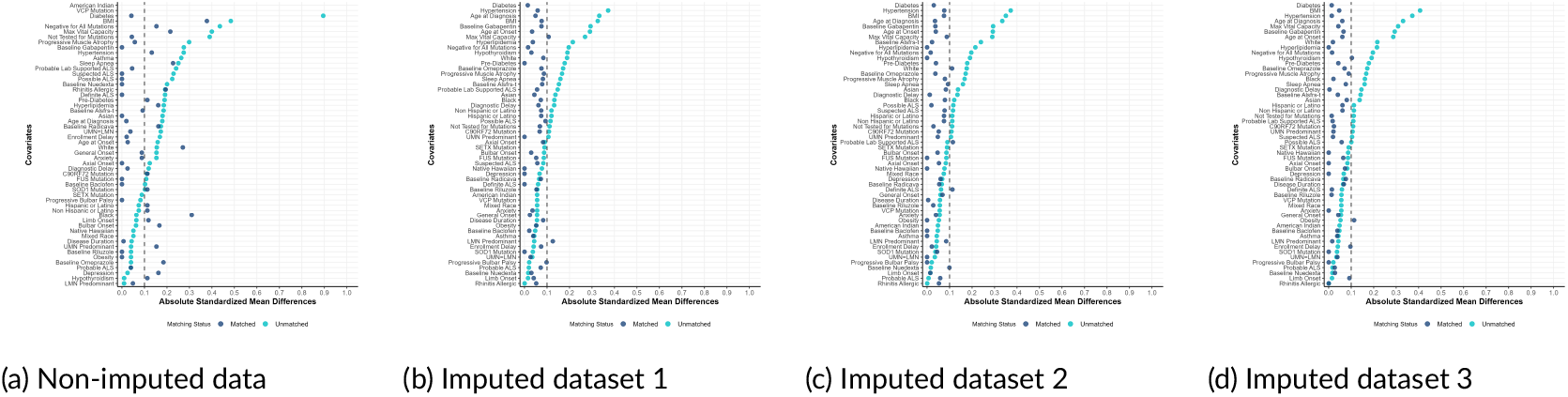
Love plots assessing balance for the metformin investigation. Each panel displays SMDs pre- and post-matching for non-imputed and imputed datasets.

Imputed dataset 1 showed estimated survival of 14.77 (metformin users) and 14.59 months (non-metformin users) for individuals never lost to follow-up (p≈0.391; N=289) and 14.75 (metformin users) and 14.68 months (non-metformin users) using principal stratification (p≈0.458; N=286). Imputed dataset 2 showed 14.87 (metformin users) and 14.43 months (non-metformin users) for those never lost to follow-up (p≈0.257; N=279) and 14.85 (metformin users) and 14.58 months (non-metformin users) using principal stratification (p≈0.351; N=275). Imputed dataset 3 showed 14.57 (metformin users) and 13.97 months (non-metformin users) for those never lost to follow-up (p≈0.191; N=301) and 14.55 (metformin users) and 13.97 months (non-metformin users) using principal stratification (p≈0.201; N=300).

Imputed matched dataset 1 exhibited the largest remaining covariate imbalance post-matching, particularly in the lower motor neuron predominant phenotype. Imputed matched dataset 1 yielded the smallest observed differences in estimated survival across both dropout strategies, suggesting that residual confounding related to phenotype may have attributed the estimated causal effect. The null randomization distributions in the metformin investigations appeared to follow gaussian distributions and centered at zero and can be found in the Appendix.

**TABLE 3.** Results of the metformin investigation. Estimated 18-month restricted mean survival time in months using 100,000 randomization iterations.

| Dataset (N Matched) | Dropout Strategy | Metformin RMST (months) | Non-metformin RMST (months) | ΔRMST | P-value |
| --- | --- | --- | --- | --- | --- |
| First imputed (N=300) | Exclude (N=289) | 14.77 | 14.59 | 0.18 | 0.391 |
|  | Never lost to follow-up (N=286) | 14.75 | 14.68 | 0.07 | 0.458 |
| Second imputed (N=292) | Exclude (N=279) | 14.87 | 14.43 | 0.44 | 0.257 |
|  | Never lost to follow-up (N=275) | 14.85 | 14.58 | 0.27 | 0.351 |
| Third imputed (N=316) | Exclude (N=301) | 14.57 | 13.97 | 0.60 | 0.191 |
|  | Never lost to follow-up (N=300) | 14.55 | 13.97 | 0.58 | 0.201 |

## 4 DISCUSSION

We undertake the baseline function and sex investigations to illustrate the framework. When evaluating the association between ALSFRS-R and survival, we rejected the null hypothesis. This finding is consistent with prior works which have demonstrated that higher baseline function is associated with extended survival in ALS (Engelberg-Cook *et al*., 2024; Daghlas and Govindarajan, 2021) and provides clarity with limited influences from confounders, demonstrating the ability of the framework to detect established relationships.

In all analyses investigating sex, we could not reject the sharp null hypothesis and have insufficient evidence against no association of sex on 18-month survival. Prior studies similarly did not identify sex as associated with survival (Knibb *et al*., 2016; Su *et al*., 2021). However, one study reported an estimated survival benefit in men with a p-value of 0.09 (Knibb *et al*., 2016) and in line with guidance from the American Statistical Association may suggest potential evidence of an association (Wasserstein *et al*., 2019). An analysis examining sex and survival found that men had shorter survival than women, though the study only considered age at onset, site of onset, diagnostic delay, cognitive status, forced vital capacity, age at diagnosis, mean monthly weight decline, and ALSFRS-R decline per month as potential confounders (Grassano *et al*., 2024). Our study accounted for additional confounders, including genetic variants and El Escorial diagnosis which are established prognostic factors (Westeneng *et al*., 2018a). Additionally, the prior study relied on asymptotic tests, while we use randomization-based inference to avoid distributional assumptions.

To our best of our knowledge, our work is the first to evaluate the causal effects of metformin in a broad population of people living with ALS. Estimated survival was larger in metformin than non-metformin user groups. However, we failed to reject the sharp null hypothesis in the metformin investigation and did not have sufficient evidence against no effect of metformin on 18-month survival. These findings are limited by relatively small sample sizes and the absence of sufficient data on medication timing and dosage. The ALS NHS remains actively recruiting and future work should consider dose-dependent effects in larger cohorts when more data is collected. A clinical trial of metformin in people living with *C9orf72*-ALS (ClinicalTrials.gov, 2025c) recruited participants from the ALS NHS. This overlap may introduce effects from study participation and is common in observational analyses where participants may be enrolled in multiple studies simultaneously.

Although we apply the RCM in a causal pipeline (Bind and Rubin, 2017) to evaluate three hypotheses in ALS, the framework is broadly applicable to other questions and disease areas. The approach can be used for investigations where randomization may not feasible as seen with baseline function and sex. Further, using observational data helps minimize burden of additional exposures and addresses ethical concerns of placebos. The use of randomization-based inference may be more suitable when working with small cohorts or heterogeneous datasets. By constructing a null randomization distribution from observed data, the randomization-based inference does not rely on distributional assumptions such as that of normality. Some have noted that significance tests that depend on irrelevant distributional assumptions may yield answers that are not well aligned with the question of interests, an issue randomization-based inference can address (Bind and Rubin, 2020). Notably, under randomization-based inference, the sharp null hypothesis is either rejected or accepted, while no direct conclusions are drawn about the alternative hypothesis.

A limitation of this study was inadequate balance to conduct subgroup analysis. We would have liked to conduct additional analyses on subgroups of ALS to assess pre-clinical findings that suggest differences in metformin efficacy across SOD1 and *C9orf72* genetic variants and sex, but were unable to due to small, imbalanced cohorts of subgroups. Future work should consider evaluating metformin across genetic variants in people living with ALS and ALS phenotypes (Meyer *et al*., 2025). Study dropout is another concern in ALS research, where attrition rates were an estimated average 33% ± 19% across 49 ALS studies (Westeneng *et al*., 2018a). Dropout threatens to introduce selection bias, particularly when data are not missing at random. Although principal stratification may help minimize exposure-related biases in such scenarios, it requires a thorough understanding of the influences of dropout for accurate modeling. Few studies have investigated participant-level dropout mechanisms and remain limited to broad analyses over interventional and observational studies (Sznajder *et al*., 2023; Atassi *et al*., 2013). Observational studies typically employ longer follow-up periods which may involve distinct dropout mechanisms, underscoring the need for further research on dropout mechanisms in ALS and observational studies. Additionally, for both randomized and observational studies, the reliability of results in ALS studies continues to be limited by outcome measures. Although several candidate biofluid, neurophysiological, and neuroimaging biomarkers have been developed to measure progression, they exhibit limited sensitivity to longitudinal changes (Kiernan *et al*., 2021). Neurofilament light chain (NfL), a marker of axonal degeneration measured in cerebrospinal fluid and plasma, has received wide attention in recent years and provides additional predictive value for disease progression and survival, yet it still does not fully explain disease heterogeneity (Benatar *et al*., 2024). Outcome and biomarker development continues to be a critical priority to more reliably evaluate treatments in ALS.

## 5 ACKNOWLEDGMENTS

The project described was supported by funding from the ALS Association, ALS Finding A Cure Foundation, the ALS and O’Hagen ALS-PLS funds of the University of Minnesota Foundation, the ALS Hope Foundation, Biogen, Harper’s Hope Fund, the Harry J Hoenselaar Endowment, the Les Turner ALS Foundation, Mitsubishi Tanabe Pharma America and by Grant Number RO1-FD007630 from FDA’s Office of Orphan Products Development. Its contents are solely the responsibility of the authors and do not necessarily represent the official views of the FDA nor FDA’s Office of Orphan Products Development. We thank Eric Macklin and Keith Barnatchez for their review of the analysis methodology.

## 6 AUTHOR CONTRIBUTIONS

J.A. Geller: drafting/revision of the manuscript for content, including medical writing for content; major role in the acquisition of data; study concept or design; analysis or interpretation of data. A. Berger: drafting/revision of the manuscript for content, including medical writing for content; major role in the acquisition of data; study concept or design; analysis or interpretation of data. M. Locatelli: drafting/revision of the manuscript for content, including medical writing for content; study concept or design. A.V. Sherman: drafting/revision of the manuscript for content, including medical writing for content; study concept or design; analysis or interpretation of data. M.A. Bind: drafting/revision of the manuscript for content, including medical writing for content; study concept or design; analysis or interpretation of data. ALS NHC Investigators: data collection, revision of the manuscript for content, including medical writing for content.

