## Supplementary Information for "Evaluating Metformin Efficacy in ALS Using Real-World Data: A Causal Inference Approach"

### Supporting Information

#### | Propensity Score Matching Implementation Details

In the function investigation, the following variables were used for propensity score estimation ($X_i$) for non-imputed and imputed datasets: sex, age at diagnosis, age at onset, race (Asian, Black), ethnicity (Hispanic or Latino), BMI, El Escorial diagnosis (possible ALS, probable ALS, probable lab supported ALS, suspected ALS), phenotype (lower motor neuron predominant, progressive bulbar palsy, upper motor neuron predominant), site of onset (axial, bulbar, general), diagnostic delay, enrollment delay, maximum vital capacity, genetic mutation status (*C9orf72*, *SOD1*, *VCP*, negative for all mutations), baseline medications (radicava, gabapentin, baclofen, nuedexta), and conditions (hypertension, depression, sleep apnea, anxiety, obesity, hypothyroidism, asthma, allergic rhinitis). The covariates for the non-imputed dataset also included *FUS* mutation, hyperlipidemia, and Native Hawaiian. Imputed dataset 1 also included riluzole, imputed dataset 2 included *SETX* mutation and riluzole, and imputed dataset 3 included *SETX* mutation and omeprazole. For all datasets in the function investigation, the caliper selected was 0.2 standard deviations.

The sex investigation included the following covariates for propensity score estimation for non-imputed and imputed datasets: age at diagnosis, age at onset, race (Asian, Native Hawaiian), BMI, El Escorial diagnosis (possible ALS, probable ALS, probable lab supported ALS), phenotype (lower motor neuron predominant, progressive bulbar palsy, progressive muscle atrophy, upper motor neuron predominant), site of onset (axial, bulbar, general), enrollment delay, genetic mutation status (*FUS*, *SOD1*, *SETX*), ALSFRS-R, baseline medications (radicava, gabapentin, omperazole), and conditions (hypertension, depression, sleep apnea, anxiety, obesity, hypothyroidism, asthma, allergic rhinitis). The covariates for non-imputed and imputed dataset 1 also included Black and nuedexta. Calipers used for non-imputed and imputed datasets were 0.5 and 0.3 respectively.

For the metformin investigation, covariates used for estimating propensity scores included the following variables for all imputed datasets: age at diagnosis, race (Asian, Black, Native Hawaiian, American Indian, Mixed Race), ethnicity (Hispanic or Latino), BMI, El Escorial diagnosis (possible ALS, probable ALS, probable lab supported ALS, suspected ALS), phenotype (lower motor neuron predominant, progressive bulbar palsy, progressive muscle atrophy, upper motor neuron predominant), site of onset (axial, bulbar, general), diagnostic delay, enrollment delay, disease duration, maximum vital capacity, genetic mutation status (*C9orf72*, *FUS*, *SETX*, *SOD1*, *VCP*, negative for all mutations), ALSFRS-R, medications (riluzole, radicava, gabapentin, baclofen, omeprazole), and conditions (pre-diabetes, diabetes, hypertension, hyperlipidemia, depression, sleep apnea, anxiety, obesity, hypothyroidism). Imputed dataset 1 also included not tested for mutations for propensity score estimation. Calipers used for non-imputed and imputed datasets 1 and 3 were 0.25 and the caliper for imputed dataset 2 was 0.3.

### Characteristics of Matched Participants

**TABLE S1 Characteristics of matched participants in the function investigation using non-imputed and imputed data.**

| | Non-imputed | | Imputed Dataset 1 | | Imputed Dataset 2 | | Imputed Dataset 3 | |
|---|---|---|---|---|---|---|---|---|
| | High Func. | Low Func. | High Func. | Low Func. | High Func. | Low Func. | High Func. | Low Func. |
| **Sex** | | | | | | | | |
| Female | 104 (43.3%) | 103 (42.9%) | 334 (41.6%) | 344 (42.8%) | 330 (41.0%) | 344 (42.7%) | 365 (42.1%) | 352 (40.6%) |
| Male | 136 (56.7%) | 137 (57.1%) | 469 (58.4%) | 459 (57.2%) | 475 (59.0%) | 461 (57.3%) | 503 (57.9%) | 516 (59.4%) |
| **Age at Onset** | | | | | | | | |
| Mean (SD) | 62.4 (10.5) | 62.6 (11.9) | 61.1 (10.7) | 61.6 (11.9) | 60.7 (11.0) | 61.7 (11.6) | 60.9 (11.1) | 61.3 (11.7) |
| Median [Min, Max] | 62.0 [29.0, 86.0] | 64.0 [23.0, 89.0] | 62.0 [25.0, 88.0] | 63.0 [20.0, 92.0] | 61.0 [20.0, 88.0] | 63.0 [20.0, 92.0] | 62.0 [12.0, 85.0] | 62.0 [20.0, 92.0] |
| **Age at Diagnosis** | | | | | | | | |
| Mean (SD) | 63.6 (10.4) | 63.9 (11.9) | 62.6 (10.5) | 63.1 (11.9) | 62.1 (10.9) | 63.2 (11.6) | 62.4 (10.9) | 62.8 (11.7) |
| Median [Min, Max] | 63.5 [37.0, 86.0] | 64.0 [24.0, 90.0] | 63.0 [27.0, 89.0] | 64.0 [20.0, 93.0] | 63.0 [21.0, 89.0] | 64.0 [20.0, 93.0] | 63.0 [21.0, 86.0] | 63.0 [20.0, 93.0] |
| **Race** | | | | | | | | |
| American Indian | - | - | | - | 3 (0.4%) | - | 2 (0.2%) | - |
| Asian | 3 (1.3%) | 3 (1.3%) | 16 (2.0%) | 9 (1.1%) | 11 (1.4%) | 9 (1.1%) | 13 (1.5%) | 12 (1.4%) |
| Black | 12 (5.0%) | 10 (4.2%) | 38 (4.7%) | 36 (4.5%) | 31 (3.9%) | 28 (3.5%) | 36 (4.1%) | 33 (3.8%) |
| Mixed Race | - | - | 3 (0.4%) | - | 3 (0.4%) | - | 4 (0.5%) | - |
| Native Hawaiian | - | - | - | 3 (0.4%) | - | 4 (0.5%) | - | 3 (0.3%) |
| White | 225 (93.8%) | 227 (94.6%) | 746 (92.9%) | 755 (94.0%) | 757 (94.0%) | 764 (94.9%) | 813 (93.7%) | 820 (94.5%) |
| **Ethnicity** | | | | | | | | |
| Hispanic or Latino | 4 (1.7%) | 4 (1.7%) | 15 (1.9%) | 13 (1.6%) | 15 (1.9%) | 17 (2.1%) | 15 (1.7%) | 15 (1.7%) |
| Non Hispanic or Latino | 236 (98.3%) | 236 (98.3%) | 788 (98.1%) | 790 (98.4%) | 790 (98.1%) | 788 (97.9%) | 853 (98.3%) | 853 (98.3%) |
| **BMI** | | | | | | | | |
| Mean (SD) | 26.3 (4.93) | 25.9 (5.87) | 26.2 (5.38) | 26.0 (6.10) | 26.4 (6.02) | 26.3 (6.46) | 26.6 (6.61) | 26.2 (6.99) |
| Median [Min, Max] | 25.9 [16.6, 51.0] | 25.0 [14.3, 45.9] | 25.5 [2.10, 51.0] | 25.0 [14.9, 89.5] | 25.6 [2.10, 85.4] | 25.1 [12.0, 96.5] | 25.5 [2.10, 89.5] | 25.0 [14.3, 96.5] |
| **El Escorial** | | | | | | | | |
| Definite ALS | 70 (29.2%) | 75 (31.3%) | 242 (30.1%) | 244 (30.4%) | 229 (28.4%) | 245 (30.4%) | 245 (28.2%) | 258 (29.7%) |
| Possible ALS | 29 (12.1%) | 31 (12.9%) | 129 (16.1%) | 111 (13.8%) | 128 (15.9%) | 118 (14.7%) | 136 (15.7%) | 126 (14.5%) |
| Probable ALS | 87 (36.3%) | 83 (34.6%) | 259 (32.3%) | 276 (34.4%) | 265 (32.9%) | 274 (34.0%) | 296 (34.1%) | 295 (34.0%) |
| Probable Lab Supported ALS | 31 (12.9%) | 26 (10.8%) | 98 (12.2%) | 93 (11.6%) | 107 (13.3%) | 101 (12.5%) | 113 (13.0%) | 102 (11.8%) |
| Suspected ALS | 23 (9.6%) | 25 (10.4%) | 75 (9.3%) | 79 (9.8%) | 76 (9.4%) | 67 (8.3%) | 78 (9.0%) | 87 (10.0%) |
| **Phenotype** | | | | | | | | |
| Lower Motor Neuron Predominant | 37 (15.4%) | 39 (16.3%) | 146 (18.2%) | 148 (18.4%) | 156 (19.4%) | 161 (20.0%) | 144 (16.6%) | 160 (18.4%) |
| Progressive Bulbar Palsy | 12 (5.0%) | 7 (2.9%) | 38 (4.7%) | 35 (4.4%) | 42 (5.2%) | 35 (4.3%) | 44 (5.1%) | 39 (4.5%) |
| Progressive Muscle Atrophy | 13 (5.4%) | 11 (4.6%) | 33 (4.1%) | 22 (2.7%) | 35 (4.3%) | 25 (3.1%) | 40 (4.6%) | 26 (3.0%) |
| Upper Motor Neuron Predominant | 28 (11.7%) | 29 (12.1%) | 90 (11.2%) | 87 (10.8%) | 89 (11.1%) | 88 (10.9%) | 103 (11.9%) | 94 (10.8%) |
| Equal Upper & Lower Motor Neuron Involvement | 150 (62.5%) | 154 (64.2%) | 496 (61.8%) | 511 (63.6%) | 483 (60.0%) | 496 (61.6%) | 537 (61.9%) | 549 (63.2%) |
| **Site of Onset** | | | | | | | | |
| Axial | 9 (3.8%) | 11 (4.6%) | 20 (2.5%) | 22 (2.7%) | 21 (2.6%) | 22 (2.7%) | 20 (2.3%) | 26 (3.0%) |
| Bulbar | 60 (25.0%) | 60 (25.0%) | 198 (24.7%) | 197 (24.5%) | 201 (25.0%) | 195 (24.2%) | 219 (25.2%) | 215 (24.8%) |
| General | 23 (9.6%) | 21 (8.8%) | 76 (9.5%) | 81 (10.1%) | 72 (8.9%) | 82 (10.2%) | 84 (9.7%) | 90 (10.4%) |
| Limb | 148 (61.7%) | 148 (61.7%) | 509 (63.4%) | 503 (62.6%) | 511 (63.5%) | 506 (62.9%) | 545 (62.8%) | 537 (61.9%) |
| **Diagnostic Delay** | | | | | | | | |
| Mean (SD) | 15.4 (16.6) | 16.3 (13.7) | 17.1 (20.2) | 17.2 (22.2) | 16.8 (21.2) | 16.4 (15.7) | 16.9 (22.3) | 17.4 (21.5) |
| Median [Min, Max] | 11 [1, 150] | 12 [0, 79] | 11 [0, 240] | 12 [0, 391] | 11 [0, 247] | 12 [0, 136] | 11 [0, 241] | 12 [0, 391] |
| **Enrollment Delay** | | | | | | | | |
| Mean (SD) | 5.39 (24.9) | 6.13 (12.3) | 6.01 (20.8) | 7.22 (18.8) | 5.62 (18.9) | 6.05 (15.6) | 5.52 (24.3) | 6.81 (20.2) |
| Median [Min, Max] | 0 [0, 349] | 1 [0, 90] | 0 [0, 349] | 1 [0, 286] | 0 [0, 349] | 1 [0, 286] | 0 [0, 483] | 1 [0, 306] |
| **Disease Duration** | | | | | | | | |
| Mean (SD) | 20.8 (35.3) | 22.4 (20.0) | 23.5 (32.1) | 24.7 (30.2) | 22.7 (31.3) | 22.8 (22.5) | 22.7 (35.6) | 24.5 (30.2) |
| Median [Min, Max] | 13 [2, 457] | 16 [3, 134] | 14 [2, 457] | 16 [2, 391] | 13 [0, 457] | 16 [2, 298] | 13 [0, 531] | 16 [2, 391] |
| **Maximum Vital Capacity** | | | | | | | | |
| Mean (SD) | 76.5 (18.2) | 75.1 (18.4) | 79.3 (18.8) | 77.3 (17.5) | 78.9 (18.9) | 77.4 (17.2) | 78.2 (19.7) | 76.5 (18.6) |
| Median [Min, Max] | 79.0 [7.00, 115] | 78.0 [8.00, 119] | 82.0 [7.00, 132] | 80.0 [17.0, 131] | 82.0 [6.00, 140] | 80.0 [19.0, 131] | 81.0 [6.00, 140] | 79.0 [5.00, 134] |
| **Genetic Mutation Status** | | | | | | | | |
| C9orf72 | 12 (5.0%) | 12 (5.0%) | 41 (5.1%) | 41 (5.1%) | 44 (5.5%) | 34 (4.2%) | 40 (4.6%) | 43 (5.0%) |
| FUS | 1 (0.4%) | 1 (0.4%) | 5 (0.6%) | 3 (0.4%) | 4 (0.5%) | 3 (0.4%) | 3 (0.3%) | 7 (0.8%) |
| Negative for All Mutations | 76 (31.7%) | 70 (29.2%) | 245 (30.5%) | 249 (31.0%) | 255 (31.7%) | 258 (32.0%) | 266 (30.6%) | 274 (31.6%) |
| Not Tested for Mutations | 145 (60.4%) | 153 (63.8%) | 491 (61.1%) | 489 (60.9%) | 478 (59.4%) | 491 (61.0%) | 534 (61.5%) | 525 (60.5%) |
| SETX | 1 (0.4%) | 1 (0.4%) | 5 (0.6%) | 3 (0.4%) | 3 (0.4%) | 4 (0.5%) | 4 (0.5%) | 3 (0.3%) |
| SOD1 | 5 (2.1%) | 3 (1.3%) | 15 (1.9%) | 16 (2.0%) | 20 (2.5%) | 14 (1.7%) | 20 (2.3%) | 14 (1.6%) |
| VCP | - | - | 1 (0.1%) | 2 (0.2%) | 1 (0.1%) | 1 (0.1%) | 1 (0.1%) | 2 (0.2%) |
| **Baseline ALSFRS-R** | | | | | | | | |
| Mean (SD) | 40.9 (2.80) | 31.5 (4.60) | 36.6 (6.54) | 41.6 (2.59) | 36.5 (6.78) | 41.7 (2.61) | 36.5 (6.76) | 41.6 (2.55) |
| Median [Min, Max] | 41.0 [37.0, 48.0] | 33.0 [11.0, 36.0] | 37.5 [3.00, 48.0] | 41.0 [38.0, 48.0] | 37.5 [4.00, 48.0] | 42.0 [38.0, 48.0] | 37.5 [1.00, 48.0] | 41.0 [38.0, 48.0] |

**TABLE S2** Characteristics of matched participants in the sex investigation using non-imputed and imputed data.

| | Non-imputed | | Imputed Dataset 1 | | Imputed Dataset 2 | | Imputed Dataset 3 | |
|---|---|---|---|---|---|---|---|---|
| | Females | Males | Females | Males | Females | Males | Females | Males |
| **Age at Onset** | | | | | | | | |
| Mean (SD) | 62.1 (12.0) | 62.6 (11.7) | 61.7 (11.9) | 61.7 (11.1) | 61.6 (12.0) | 61.4 (11.1) | 61.5 (12.0) | 61.1 (11.3) |
| Median [Min, Max] | 64.0 [23.0, 87.0] | 64.0 [13.0, 93.0] | 63.0 [19.0, 90.0] | 62.0 [21.0, 92.0] | 63.0 [16.0, 90.0] | 62.0 [21.0, 92.0] | 63.0 [9.00, 90.0] | 62.0 [17.0, 92.0] |
| **Age at Diagnosis** | | | | | | | | |
| Mean (SD) | 63.4 (12.0) | 63.9 (11.5) | 63.1 (11.8) | 63.1 (11.1) | 63.0 (11.9) | 62.8 (11.1) | 62.9 (11.9) | 62.5 (11.2) |
| Median [Min, Max] | 65.0 [24.0, 88.0] | 65.0 [26.0, 94.0] | 64.0 [20.0, 91.0] | 64.0 [22.0, 93.0] | 64.0 [16.0, 91.0 | 63.0 [22.0, 93.0] | 64.0 [16.0, 91.0] | 64.0 [18.0, 93.0] |
| **Race** | | | | | | | | |
| American Indian | - | - | - | 3 (0.3%) | - | 3 (0.3%) | - | 4 (0.4%) |
| Asian | 8 (2.5%) | 8 (2.5%) | 20 (1.8%) | 19 (1.7%) | 19 (1.7%) | 18 (1.6%) | 19 (1.7%) | 17 (1.5%) |
| Black | 18 (5.5%) | 17 (5.2%) | 59 (5.2%) | 51 (4.5%) | 61 (5.4%) | 57 (5.0%) | 65 (5.7%) | 55 (4.8%) |
| Native Hawaiian | - | - | - | - | - | - | 3 (0.3%) | 2 (0.2%) |
| Mixed Race | - | - | 3 (0.3%) | 1 (0.1%) | 3 (0.3%) | 1 (0.1%) | 3 (0.3%) | 1 (0.1%) |
| White | 299 (92.0%) | 300 (92.3%) | 1046 (92.7%) | 1054 (93.4%) | 1054 (92.7%) | 1058 (93.1%) | 1050 (92.1%) | 1061 (93.1%) |
| **Ethnicity** | | | | | | | | |
| Hispanic or Latino | 7 (2.2%) | 7 (2.2%) | 29 (2.6%) | 20 (1.8%) | 28 (2.5%) | 17 (1.5%) | 28 (2.5%) | 21 (1.8%) |
| Non Hispanic or Latino | 318 (97.8%) | 318 (97.8%) | 1099 (97.4%) | 1108 (98.2%) | 1109 (97.5%) | 1120 (98.5%) | 1112 (97.5%) | 1119 (98.2%) |
| **BMI** | | | | | | | | |
| Mean (SD) | 26.1 (6.34) | 26.0 (4.35) | 26.2 (7.12) | 26.1 (5.54) | 26.1 (6.89) | 26.0 (5.63) | 26.0 (6.64) | 25.9 (5.58) |
| Median [Min, Max] | 24.6 [2.10, 51.0] | 25.6 [14.3, 44.5] | 25.1 [2.10, 96.5] | 25.5 [14.3, 89.5] | 24.9 [2.10, 96.5] | 25.5 [2.10, 85.4] | 24.9 [2.10, 96.5] | 25.1 [2.10, 89.5] |
| **El Escorial** | | | | | | | | |
| Definite ALS | 101 (31.1%) | 98 (30.2%) | 346 (30.7%) | 344 (30.5%) | 346 (30.4%) | 345 (30.3%) | 353 (31.0%) | 357 (31.3%) |
| Possible ALS | 44 (13.5%) | 44 (13.5%) | 177 (15.7%) | 173 (15.3%) | 178 (15.7%) | 174 (15.3%) | 182 (16.0%) | 178 (15.6%) |
| Probable ALS | 108 (33.2%) | 104 (32.0%) | 374 (33.2%) | 373 (33.1%) | 386 (33.9%) | 400 (35.2%) | 385 (33.8%) | 393 (34.5%) |
| Probable Lab Supported ALS | 51 (15.7%) | 53 (16.3%) | 156 (13.8%) | 158 (14.0%) | 153 (13.5%) | 157 (13.8%) | 149 (13.1%) | 149 (13.1%) |
| Suspected ALS | 21 (6.5%) | 26 (8.0%) | 75 (6.6%) | 80 (7.1%) | 74 (6.5%) | 61 (5.4%) | 71 (6.2%) | 63 (5.5%) |
| **Phenotype** | | | | | | | | |
| Lower Motor Neuron Predominant | 38 (11.7%) | 43 (13.2%) | 164 (14.5%) | 178 (15.8%) | 165 (14.5%) | 180 (15.8%) | 166 (14.6%) | 170 (14.9%) |
| Progressive Bulbar Palsy | 16 (4.9%) | 11 (3.4%) | 71 (6.3%) | 56 (5.0%) | 75 (6.6%) | 56 (4.9%) | 74 (6.5%) | 56 (4.9%) |
| Progressive Muscle Atrophy | 11 (3.4%) | 9 (2.8%) | 17 (1.5%) | 18 (1.6%) | 17 (1.5%) | 14 (1.2%) | 17 (1.5%) | 20 (1.8%) |
| Upper Motor Neuron Predominant | 47 (14.5%) | 42 (12.9%) | 159 (14.1%) | 145 (12.9%) | 157 (13.8%) | 140 (12.3%) | 157 (13.8%) | 145 (12.7%) |
| Equal Upper & Lower Motor Neuron Involvement | 213 (65.5%) | 220 (67.7%) | 717 (63.6%) | 731 (64.8%) | 723 (63.6%) | 747 (65.7%) | 726 (63.7%) | 749 (65.7%) |
| **Site of Onset** | | | | | | | | |
| Axial | 9 (2.8%) | 7 (2.2%) | 20 (1.8%) | 19 (1.7%) | 20 (1.8%) | 15 (1.3%) | 21 (1.8%) | 18 (1.6%) |
| Bulbar | 88 (27.1%) | 85 (26.2%) | 327 (29.0%) | 289 (25.6%) | 332 (29.2%) | 285 (25.1%) | 331 (29.0%) | 291 (25.5%) |
| General | 27 (8.3%) | 33 (10.2%) | 121 (10.7%) | 128 (11.3%) | 121 (10.6%) | 131 (11.5%) | 124 (10.9%) | 127 (11.1%) |
| Limb | 201 (61.8%) | 200 (61.5%) | 660 (58.5%) | 692 (61.3%) | 664 (58.4%) | 706 (62.1%) | 664 (58.2%) | 704 (61.8%) |
| **Diagnostic Delay** | | | | | | | | |
| Mean (SD) | 16.0 (14.2) | 16.9 (20.8) | 16.3 (16.1) | 16.6 (20.0) | 16.4 (16.3) | 15.6 (16.4) | 16.7 (16.9) | 16.8 (20.6) |
| Median [Min, Max] | 11 [1, 81] | 11 [0, 241] | 11 [0, 146] | 11 [0, 264] | 11 [0, 146] | 11 [0, 150] | 11 [0, 146] | 11 [0, 264] |
| **Enrollment Delay** | | | | | | | | |
| Mean (SD) | 6.88 (23.2) | 5.78 (14.6) | 9.47 (30.4) | 8.81 (28.3) | 9.90 (33.1) | 9.02 (28.3) | 9.75 (32.9) | 9.01 (26.8) |
| Median [Min, Max] | 1 [0, 258] | 1 [0, 117] | 1 [0, 483] | 1 [0, 412] | 1 [0, 483] | 1 [0, 412] | 1 [0, 483] | 1 [0, 412] |
| **Disease Duration** | | | | | | | | |
| Mean (SD) | 22.9 (29.5) | 22.7 (27.3) | 26.1 (37.7) | 25.7 (36.7) | 26.6 (39.8) | 24.9 (34.5) | 26.7 (40.7) | 26.1 (35.5) |
| Median [Min, Max] | 13 [2, 298] | 14 [2, 241] | 14 [2, 531] | 15 [0, 457] | 15 [2, 531] | 15 [0, 457] | 15 [2, 531] | 15 [2, 412] |
| **Maximum Vital Capacity** | | | | | | | | |
| Mean (SD) | 75.4 (21.8) | 75.8 (21.3) | 72.1 (23.7) | 73.7 (22.8) | 72.1 (23.6) | 73.4 (23.8) | 72.2 (24.0) | 72.9 (23.6) |
| Median [Min, Max] | 80.0 [8.00, 129] | 80.0 [7.00, 145] | 76.0 [6.00, 130] | 78.5 [5.00, 145] | 76.0 [6.00, 130] | 79.0 [5.00, 147] | 76.0 [5.00, 134] | 78.0 [5.00, 147] |
| **Genetic Mutation Status** | | | | | | | | |
| C9orf72 | 21 (6.5%) | 14 (4.3%) | 68 (6.0%) | 46 (4.1%) | 68 (6.0%) | 49 (4.3%) | 69 (6.1%) | 49 (4.3%) |
| FUS | - | 1 (0.3%) | 5 (0.4%) | 5 (0.4%) | 5 (0.4%) | 6 (0.5%) | 5 (0.4%) | 6 (0.5%) |
| Negative for All Mutations | 88 (27.1%) | 103 (31.7%) | 356 (31.6%) | 357 (31.6%) | 359 (31.6%) | 349 (30.7%) | 358 (31.4%) | 365 (32.0%) |
| Not Tested for Mutations | 208 (64.0%) | 199 (61.2%) | 659 (58.4%) | 688 (61.0%) | 668 (58.8%) | 704 (61.9%) | 670 (58.8%) | 691 (60.6%) |
| SETX | 1 (0.3%) | 1 (0.3%) | 5 (0.4%) | 3 (0.3%) | 4 (0.4%) | 3 (0.3%) | 4 (0.4%) | 3 (0.3%) |
| SOD1 | 7 (2.2%) | 7 (2.2%) | 32 (2.8%) | 28 (2.5%) | 30 (2.6%) | 25 (2.2%) | 31 (2.7%) | 25 (2.2%) |
| VCP | - | - | 3 (0.3%) | 1 (0.1%) | 3 (0.3%) | 1 (0.1%) | 3 (0.3%) | 1 (0.1%) |
| **Baseline ALSFRS-R** | | | | | | | | |
| Mean (SD) | 36.1 (7.17) | 36.5 (6.66) | 34.3 (8.61) | 34.8 (8.48) | 34.2 (8.60) | 34.8 (8.51) | 34.4 (8.64) | 34.9 (8.37) |
| Median [Min, Max] | 37.0 [11.0, 47.0] | 37.0 [16.0, 48.0] | 36.0 [1.00, 48.0] | 36.0 [2.00, 48.0] | 36.0 [1.00, 48.0] | 36.0 [1.00, 48.0] | 36.0 [1.00, 48.0] | 36.0 [1.00, 48.0] |

**TABLE S3** Characteristics of matched participants in the metformin investigation using imputed data.

| | Imputed Dataset 1 | | Imputed Dataset 2 | | Imputed Dataset 3 | |
|---|---|---|---|---|---|---|
| | Metformin | Non-metformin | Metformin | Non-metformin | Metformin | Non-metformin |
| **Sex** | | | | | | |
| Female | 58 (38.7%) | 63 (42.0%) | 54 (37.0%) | 80 (54.8%) | 59 (37.3%) | 66 (41.8%) |
| Male | 92 (61.3%) | 87 (58.0%) | 92 (63.0%) | 66 (45.2%) | 99 (62.7%) | 92 (58.2%) |
| **Age at Onset** | | | | | | |
| Mean (SD) | 63.2 (9.39) | 62.8 (10.9) | 63.2 (9.52) | 63.6 (10.9) | 63.3 (9.39) | 62.6 (11.5) |
| Median [Min, Max] | 64.0 [35.0, 85.0] | 64.0 [30.0, 88.0] | 64.0 [35.0, 85.0] | 64.0 [36.0, 90.0] | 64.0 [35.0, 85.0] | 64.0 [30.0, 88.0] |
| **Age at Diagnosis** | | | | | | |
| Mean (SD) | 64.9 (8.88) | 64.4 (10.9) | 65.0 (9.04) | 65.3 (10.8) | 65.2 (9.00) | 64.5 (11.3) |
| Median [Min, Max] | 65.0 [36.0, 85.0] | 66.0 [31.0, 89.0] | 65.0 [36.0, 86.0] | 66.0 [39.0, 91.0] | 66.0 [36.0, 86.0] | 66.0 [31.0, 89.0] |
| **Race** | | | | | | |
| American Indian | - | - | - | - | - | - |
| Asian | 4 (2.7%) | 3 (2.0%) | 5 (3.4%) | 3 (2.1%) | 5 (3.2%) | 3 (1.9%) |
| Black | 14 (9.3%) | 11 (7.3%) | 12 (8.2%) | 9 (6.2%) | 15 (9.5%) | 16 (10.1%) |
| Mixed Race | - | - | - | - | - | - |
| Native Hawaiian | 1 (0.7%) | 1 (0.7%) | 1 (0.7%) | 1 (0.7%) | 1 (0.6%) | 1 (0.6%) |
| White | 131 (87.3%) | 135 (90.0%) | 128 (87.7%) | 133 (91.1%) | 137 (86.7%) | 138 (87.3%) |
| **Ethnicity** | | | | | | |
| Hispanic or Latino | 6 (4.0%) | 4 (2.7%) | 6 (4.1%) | 4 (2.7%) | 6 (3.8%) | 8 (5.1%) |
| Non Hispanic or Latino | 144 (96.0%) | 146 (97.3%) | 140 (95.9%) | 142 (97.3%) | 152 (96.2%) | 150 (94.9%) |
| **BMI** | | | | | | |
| Mean (SD) | 27.9 (5.84) | 28.5 (9.43) | 28.0 (5.91) | 28.6 (9.35) | 28.6 (8.18) | 29.1 (11.2) |
| Median [Min, Max] | 27.2 [15.4, 47.0] | 27.2 [17.6, 96.5] | 27.0 [15.9, 44.5] | 27.0 [17.0, 96.5] | 27.2 [12.0, 89.5] | 27.2 [16.0, 96.5] |
| **El Escorial** | | | | | | |
| Definite ALS | 37 (24.7%) | 37 (24.7%) | 38 (26.0%) | 31 (21.2%) | 44 (27.8%) | 43 (27.7%) |
| Possible ALS | 20 (13.3%) | 25 (16.7%) | 21 (14.4%) | 22 (15.1%) | 21 (13.3%) | 18 (11.4%) |
| Probable ALS | 49 (32.7%) | 44 (29.3%) | 49 (33.6%) | 45 (30.8%) | 50 (31.6%) | 52 (32.9%) |
| Probable Lab Supported ALS | 24 (16.0%) | 21 (14.0%) | 19 (13.0%) | 25 (17.1%) | 24 (15.2%) | 25 (15.8%) |
| Suspected ALS | 20 (13.3%) | 23 (15.3%) | 19 (13.0%) | 23 (15.8%) | 19 (12.0%) | 20 (12.7%) |
| **Phenotype** | | | | | | |
| Lower Motor Neuron Predominant | 31 (20.7%) | 39 (26.0%) | 32 (21.9%) | 27 (18.5%) | 33 (20.9%) | 32 (20.3%) |
| Progressive Bulbar Palsy | 8 (5.3%) | 5 (3.3%) | 7 (4.8%) | 7 (4.8%) | 8 (5.1%) | 8 (5.1%) |
| Progressive Muscle Atrophy | 10 (6.7%) | 7 (4.7%) | 9 (6.2%) | 12 (8.2%) | 9 (5.7%) | 6 (3.8%) |
| Upper Motor Neuron Predominant | 13 (8.7%) | 13 (8.7%) | 12 (8.2%) | 14 (9.6%) | 15 (9.5%) | 16 (10.1%) |
| Equal Upper & Lower Motor Neuron Involvement | 88 (58.7%) | 86 (57.3%) | 86 (58.9%) | 86 (58.9%) | 93 (58.9%) | 96 (60.8%) |
| **Site of Onset** | | | | | | |
| Axial | 3 (2.0%) | 5 (3.3%) | 2 (1.4%) | 3 (2.1%) | 2 (1.3%) | 2 (1.3%) |
| Bulbar | 41 (27.3%) | 43 (28.7%) | 39 (26.7%) | 36 (24.7%) | 43 (27.2%) | 38 (24.1%) |
| General | 13 (8.7%) | 12 (8.0%) | 13 (8.9%) | 16 (11.0%) | 16 (10.1%) | 14 (8.9%) |
| Limb | 93 (62.0%) | 90 (60.0%) | 92 (63.0%) | 91 (62.3%) | 97 (61.4%) | 104 (65.8%) |
| **Diagnostic Delay** | | | | | | |
| Mean (SD) | 18.7 (25.7) | 17.4 (16.4) | 19.9 (30.5) | 19.6 (24.3) | 21.2 (30.4) | 21.3 (22.8) |
| Median [Min, Max] | 12 [0, 264] | 12 [0, 119] | 11.5 [0, 264] | 12 [0, 216] | 12 [2, 264] | 13 [0, 146] |
| **Enrollment Delay** | | | | | | |
| Mean (SD) | 8.83 (29.6) | 7.05 (17.2) | 8.95 (29.9) | 9.54 (26.2) | 8.28 (28.8) | 11.4 (34.7) |
| Median [Min, Max] | 0 [0, 201] | 0 [0, 130] | 0 [0, 201] | 0 [0, 147] | 0 [0, 201] | 1.00 [0, 306] |
| **Disease Duration** | | | | | | |
| Mean (SD) | 27.8 (45.9) | 24.6 (28.5) | 29.1 (48.8) | 29.4 (39.6) | 29.8 (47.5) | 33.0 (47.5) |
| Median [Min, Max] | 14 [2, 377] | 15 [2, 250] | 13 [2, 377] | 15 [2, 250] | 14 [2, 377] | 18 [3, 330] |
| **Maximum Vital Capacity** | | | | | | |
| Mean (SD) | 68.3 (23.3) | 65.8 (23.5) | 69.7 (22.8) | 71.7 (23.8) | 67.4 (23.0) | 68.5 (24.5) |
| Median [Min, Max] | 73.0 [8.00, 108] | 66.5 [8.00, 117] | 73.0 [9.00, 108] | 76.0 [7.00, 124] | 71.0 [21.0, 118] | 72.0 [7.00, 120] |
| **Genetic Mutation Status** | | | | | | |
| C9orf72 | 13 (8.7%) | 16 (10.7%) | 12 (8.2%) | 10 (6.8%) | 13 (8.2%) | 12 (7.6%) |
| FUS | 2 (1.3%) | 3 (2.0%) | 2 (1.4%) | 2 (1.4%) | 2 (1.3%) | 1 (0.6%) |
| Negative for All Mutations | 35 (23.3%) | 36 (24.0%) | 36 (24.7%) | 37 (25.3%) | 37 (23.4%) | 38 (24.1%) |
| Not Tested for Mutations | 95 (63.3%) | 90 (60.0%) | 92 (63.0%) | 94 (64.4%) | 101 (63.9%) | 102 (64.6%) |
| SETX | - | - | - | - | - | - |
| SOD1 | 5 (3.3%) | 5 (3.3%) | 4 (2.7%) | 3 (2.1%) | 5 (3.2%) | 5 (3.2%) |
| VCP | - | - | - | - | - | - |
| **Baseline ALSFRS-R** | | | | | | |
| Mean (SD) | 34.2 (9.73) | 34.9 (8.20) | 34.1 (9.62) | 34.3 (8.25) | 33.9 (9.08) | 33.5 (8.96) |
| Median [Min, Max] | 36.0 [4.0, 47.0] | 37.0 [10.0, 47.0] | 36.0 [1.00, 48.0] | 36.0 [2.00, 47.0] | 36.0 [1.00, 48.0] | 36.0 [6.00, 47.0] |
| **Concurrent Conditions** | | | | | | |
| Pre-Diabetes | 4 (2.7%) | 4 (2.7%) | 4 (2.7%) | 5 (3.4%) | 4 (2.5%) | 3 (1.9%) |
| Diabetes | 49 (32.7%) | 48 (32.0%) | 45 (30.8%) | 43 (29.5%) | 57 (36.1%) | 58 (36.7%) |

### | Love Plots Assessing Matching Balance for Secondary Analyses

**FIGURE S1 Love plots assessing balance for the function investigation.** Each panel displays SMDs pre- and post-matching matching for non-imputed and imputed datasets.

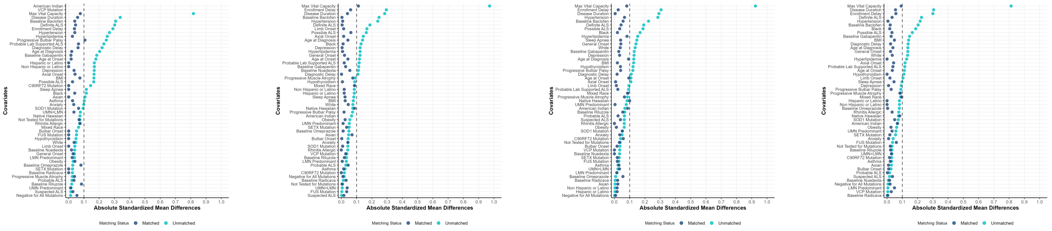


(a) Non-imputed data (b) Imputed dataset 1 (c) Imputed dataset 2 (d) Imputed dataset 3

**FIGURE S2 Love plots assessing balance for the sex investigation.** Each panel displays SMDs pre- and post-matching matching for non-imputed and imputed datasets.

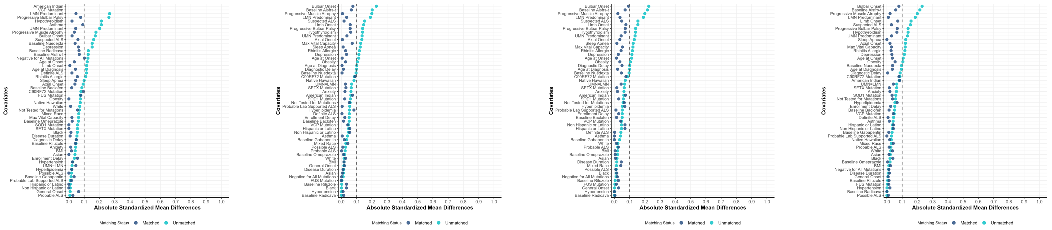


(a) Non-imputed data (b) Imputed dataset 1 (c) Imputed dataset 2 (d) Imputed dataset 3

### Null Randomization Distributions under Randomization-Based Inference

**FIGURE S3 Null randomization distributions for the function investigation showing the difference in 18-month restricted mean survival time for matched non-imputed and imputed data.** The top two rows show distributions after applying the naive dropout strategy and bottom two rows after conducting principal stratification.

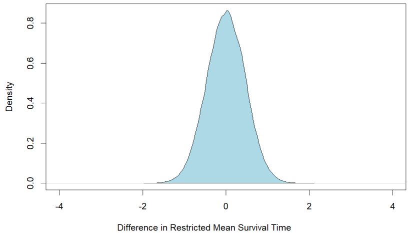


(a) Naive, non-imputed data

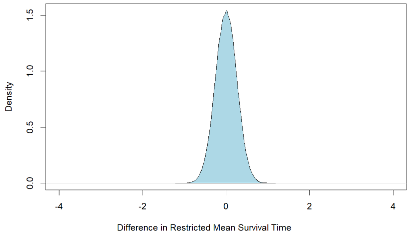


(b) Naive, imputed dataset 1

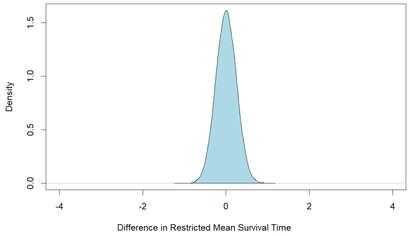


(c) Naive, imputed dataset 2

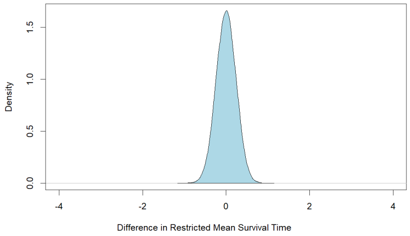


(d) Naive, imputed dataset 3

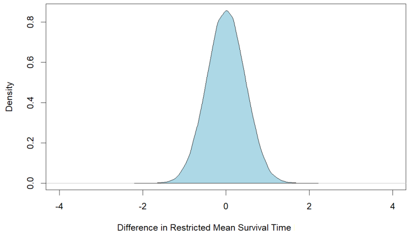


(e) Principal stratification, non-imputed data

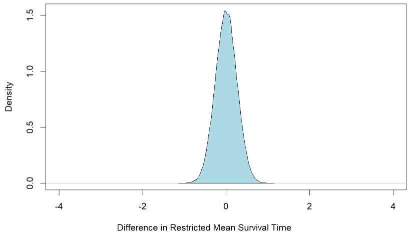


(f) Principal stratification, imputed dataset 1

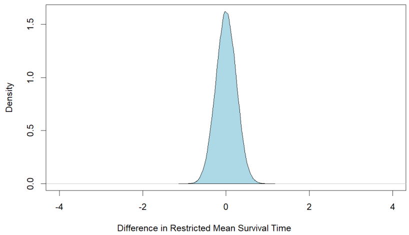


(g) Principal stratification, imputed dataset 2

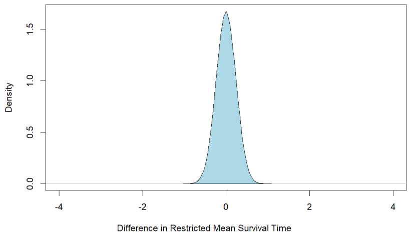


(h) Principal stratification, imputed dataset 3

**FIGURE S4 Null randomization distributions for the sex investigation showing the difference in 18-month restricted mean survival time for matched non-imputed and imputed data.** The top two rows show distributions after applying the naive dropout strategy and bottom two rows after conducting principal stratification.

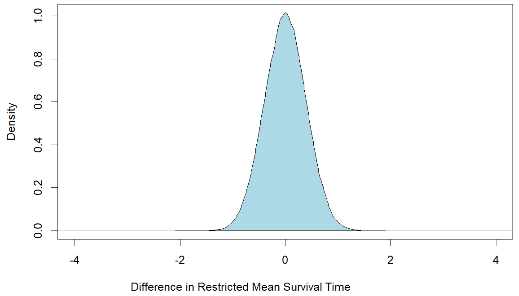


(a) Naive, non-imputed data

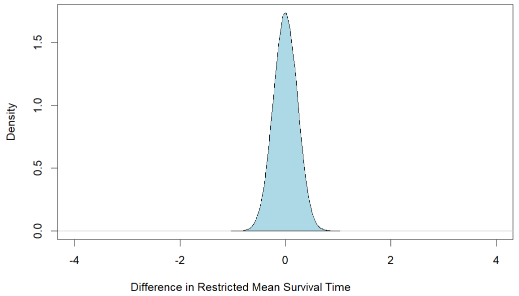


(b) Naive, imputed dataset 1

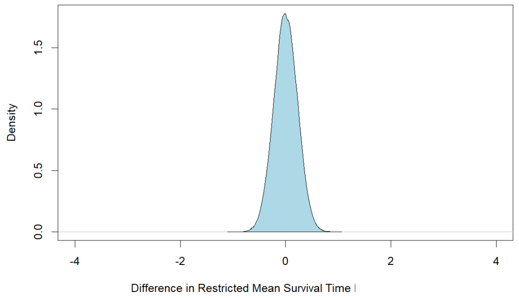


(c) Naive, imputed dataset 2

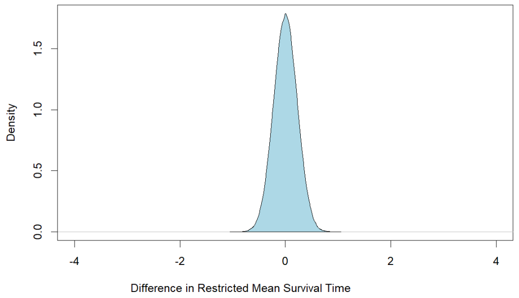


(d) Naive, imputed dataset 3

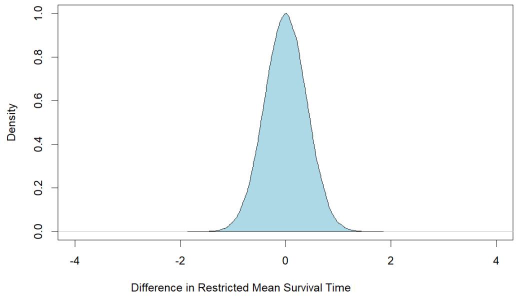


(e) Principal stratification, non-imputed data

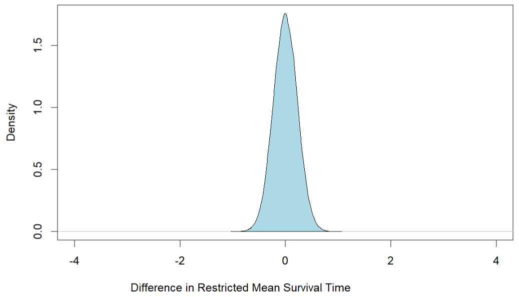


(f) Principal stratification, imputed dataset 1

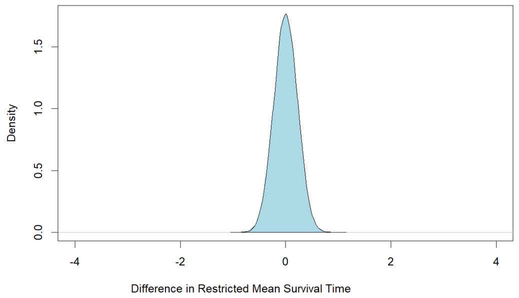


(g) Principal stratification, imputed dataset 2

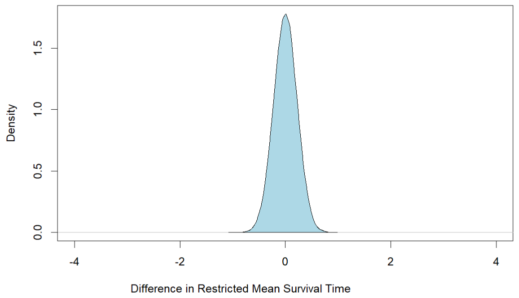


(h) Principal stratification, imputed dataset 3

**FIGURE S5 Null randomization distributions for the metformin investigation showing the difference in 18-month restricted mean survival time for matched imputed data.** Top panel shows distributions after applying the naive dropout strategy and bottom panel after conducting principal stratification.

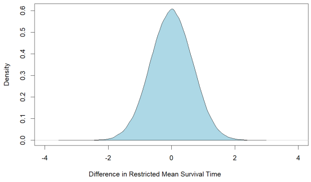


(a) Naive, imputed dataset 1

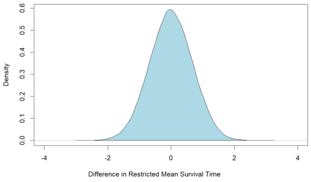


(b) Naive, imputed dataset 2

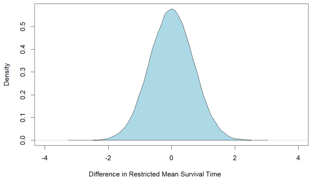


(c) Naive, imputed dataset 3

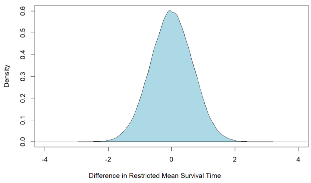


(d) Principal stratification, imputed dataset 1

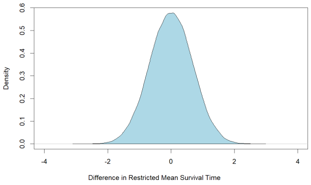


(e) Principal stratification, imputed dataset 2

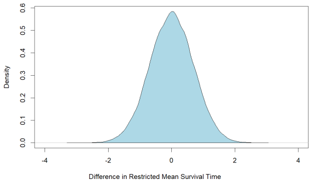


(f) Principal stratification, imputed dataset 3
